# Revealing the Infiltration: Prognostic Value of Automated Segmentation of Non-Contrast-Enhancing Tumor in Glioblastoma

**DOI:** 10.1101/2025.06.26.25327675

**Authors:** Maria Gómez-Mahiques, Carles Lopez-Mateu, F. Javier Gil-Terrón, Victor Montosa-i-Micó, Siri Fløgstad Svensson, Eduardo Erasmo Mendoza Mireles, Einar Osland Vik-Mo, Kyrre E. Emblem, Carme Balañà-Quintero, Josep Puig, Juan M. García-Gómez, Elies Fuster-Garcia

## Abstract

**Background:** Precise delineation of non-contrast-enhancing tumor (nCET) in glioblastoma (GB) is critical for maximal safe resection, yet routine imaging cannot reliably separate infiltrative tumor from vasogenic edema. The aim of this study was to develop and validate an automated method to identify nCET and assess its prognostic value.

**Methods:** Pre-operative T2-weighted and FLAIR MRI from 940 patients with newly diagnosed GB in four multicenter cohorts were analyzed. A deep-learning model segmented enhancing tumor, edema and necrosis; a non-local spatially varying finite mixture model then isolated edema subregions containing nCET. The ratio of nCET to total edema volume—the Diffuse Infiltration Index (DII)—was calculated. Associations between DII and overall survival (OS) were examined with Kaplan–Meier curves and multivariable Cox regression.

**Results:** The algorithm distinguished nCET from vasogenic edema in 97.5 % of patients, showing a mean signal-intensity gap > 5 %. Higher DII is able to stratify patients with shorter OS. In the NCT03439332 cohort, DII above the optimal threshold doubled the hazard of death (hazard ratio 2.09, 95 % confidence interval 1.34–3.25; p = 0.0012) and reduced median survival by 122 days. Significant, though smaller, effects were confirmed in GLIOCAT & BraTS (hazard ratio 1.31; p = 0.022), OUS (hazard ratio 1.28; p = 0.007) and in pooled analysis (hazard ratio 1.28; p = 0.0003). DII remained an independent predictor after adjustment for age, extent of resection and MGMT methylation.

**Conclusions:** We present a reproducible, server-hosted tool for automated nCET delineation and DII biomarker extraction that enables robust, independent prognostic stratification. It promises to guide supramaximal surgical planning and personalized neuro-oncology research and care.

**Key Points:**

‐ KP1: Robust automated MRI tool segments non-contrast-enhancing (nCET) glioblastoma.
‐ KP2: Introduced and validated the Diffuse Infiltration Index with prognostic value.
‐ KP3: nCET mapping enables RANO supramaximal resection for personalized surgery.

**Importance of the Study:** This study underscores the clinical importance of accurately delineating non-contrast-enhancing tumor (nCET) regions in glioblastoma (GB) using standard MRI. Despite their lack of contrast enhancement, nCET areas often harbor infiltrative tumor cells critical for disease progression and recurrence. By integrating deep learning segmentation with a non-local finite mixture model, we developed a reproducible, automated methodology for nCET delineation and introduced the Diffuse Infiltration Index (DII), a novel imaging biomarker. Higher DII values were independently associated with reduced overall survival across large, heterogeneous cohorts. These findings highlight the prognostic relevance of imaging-defined infiltration patterns and support the use of nCET segmentation in clinical decision-making. Importantly, this methodology aligns with and operationalizes recent RANO criteria on supramaximal resection, offering a practical, image-based tool to improve surgical planning. In doing so, our work advances efforts toward more personalized neuro-oncological care, potentially improving outcomes while minimizing functional compromise.

## Introduction

Gliomas are among the most prevalent types of primary brain tumors, accounting for over 70% of all malignant brain tumors ^1^. Glioblastoma (GB) is a particularly aggressive tumor of the central nervous system, with a median survival of only 12-14 months following diagnosis ^1^, despite standard of care that includes tumor resection followed by radiotherapy and concomitant and adjuvant cycles of temozolomide ^2 3^.

In the quest to improve the prognosis for patients with GB, different extents of surgical resection have been explored. Studies have demonstrated that supramaximal resection of a magnetic resonance imaging (MRI)-based abnormality zone beyond the contrast-enhancing tumor (CET) is associated with prolonged patient survival ^4,5^. Consequently, while this practice is currently recommended as a guideline ^6,7^, it does not explicitly define which regions should be included in the supramaximal resection, leaving it to the discretion of the neurosurgeon ^6^. Some experts suggest resecting 1-2 cm of tissue around the CET, thus targeting a part of the surrounding edematous region, which is morphologically discernible on conventional MRIs ^6^. This region is biologically heterogeneous, and the surgical approach inevitably involves removing both malignant and non-pathological tissue, which may be detrimental to the patient’s quality of life and could potentially impact their clinical outcome.

In this context, the non-contrast-enhancing tumor (nCET) region, extending beyond the contrast-enhanced areas, plays a crucial role. While challenging to delineate, it can often be identified using T2-weighted fluid-attenuated inversion recovery (FLAIR) and T2-weighted (T2), MRI ^8,9^. nCET has been described to contain higher concentration of tumor cells than CET ^10^. Incomplete resection of the nCET region has been linked to earlier tumor recurrence ^2^. In contrast, supramaximal resection, which includes the removal of both CET and part of the surrounding FLAIR hyperintensity, has been associated with improved survival. Pessina et al. ^11^ reported a median survival of 29 months for patients undergoing supramaximal resection, compared to 16 months for those with only macroscopic total resection (>90% of CET). However, a standardized, reproducible approach for defining and segmenting the nCET region has yet to be established, posing a challenge for consistent diagnosis and treatment planning ^9^. Recent reviews by the Response Assessment in Neuro-Oncology (RANO) resect group ^7^ and Annette et al.^12^ further reinforced these findings, analyzing multiple studies on the impact of resection extent in GB. The authors concluded that achieving a supramaximal resection—defined as the complete removal of CET with ≤ 5 cm^3^ of residual nCET ^7^—correlates with a better prognosis. However, they highlight the complexity of quantifying resection extent, particularly in distinguishing tumor-infiltrated tissue from vasogenic edema on T2w and FLAIR MRIs. RANO’s study underscores the absence of a standardized method to delineate nCET boundaries, stressing the risks of indiscriminately resecting vasogenic edema regions, potentially impacting the patient’s functional capacity without patient benefit.

Our study aimed to develop a precise and reproducible methodology for defining the nCET region on T2w and FLAIR MRIs. By associating the nCET definition outcomes with the overall survival of GB patients, we sought to underline the clinical relevance of accurately identifying this region. Our approach is designed to serve as a practical tool, particularly for planning supramaximal resections, ultimately providing neurosurgeons with a more informed basis for surgical planning and striving to improve prognostic outcomes for patients with GB.

## Materials and methods

### Patient cohorts

The following section presents a comprehensive overview of the datasets employed in our study. Each dataset was derived from multiple clinical institutions operating under ethically approved protocols.

#### NCT03439332 dataset

This study included data from GB patients from seven European clinical centres: Hospital Universitario de La Ribera (Alzira, Spain), Hospital de Manises (Manises, Spain), Hospital Clínic (Barcelona, Spain), Hospital Universitario Vall d’Hebron (Barcelona, Spain), Azienda Ospedaliero-Universitaria di Parma (Parma, Italy), Centre Hospitalier Universitaire de Liège (Liège, Belgium), and Oslo University Hospital – OUS (Oslo, Norway)^13^. Patients were diagnosed with GB grade IV 2016 WHO with histopathological confirmation and followed Stupp standard treatment. The extent of resection was assessed in each center by expert neurosurgeons and radiologists based on the postsurgical MRI study findings. The dataset includes comprehensive clinical, molecular, and MRI data: T1-weighted (T1), contrast-enhanced T1-weighted (T1c), T2-weighted (T2), Fluid Attenuated Inversion Recovery (T2-FLAIR), and Dynamic Susceptibility Contrast perfusion (DSC). After applying the eligibility criteria (see below), 113 patients were retained for analysis.

#### GLIOCAT dataset

This dataset consists of patients from six different institutions: Instituto Catalán de Oncología (ICO) de Badalona (Barcelona), Hospital del Mar (Barcelona), Hospital Clínic (Barcelona), ICO Hospitalet (Barcelona), ICO Girona (Girona), and Hospital Sant Pau (Barcelona). The cohort is part of the retrospective multicentric Gliocat study ^14^, derived from the Gliocat Project, and includes patients diagnosed with GB according to the 2021 WHO CNS tumor classification. All patients received standard first-line treatment, consisting of surgery followed by radiotherapy with concurrent and adjuvant temozolomide, between 2004 and 2015. Additionally, the dataset includes comprehensive clinical, molecular, and MRI (T1, T1c, T2, FLAIR and DSC) data. A total of 118 patients met the inclusion criteria.

#### BraTS dataset

A publicly available dataset from the Multimodal Brain Tumor Segmentation Challenge 2019 (BraTS 2019), organized as part of the international MICCAI 2019 conference^15–17^. This dataset comprises multi-parametric MRI (T1, T1c, T2 and FLAIR) scans and limited clinical data including age, sex, and extent of resection from patients diagnosed with GB.Specifically included in the training corpus for the BraTS 2019 Overall Survival (OS) prediction challenge. It integrates data from the 2013 BraTS dataset and additional scans from the Center for Biomedical Image Computing and Analytics (CBICA) and The Cancer Imaging Archive (TCIA). This yielded 211 eligible patients.

#### OUS dataset

he dataset has been used in a previous publication^18^ and based upon a retrospective cohort of glioblastoma patients at OUS). Since 2003, all patients undergoing first-time surgery have been prospectively registered. For our study we included all patients who (1) were diagnosed with a histopathologically verified supratentorial GB (2003–2016), GB WHO grade IV (2016– 2019), or tumors classified as gliosarcoma, giant cell GB, or epithelioid GB, according to the relevant WHO classification of tumors of the central nervous system at the time; (2) had undergone surgical resection of the tumor between 2003 and 2020 and (3) had a preoperative anatomical MRI (T1,T2 and FLAIR) scan. The final cohort comprised 498 eligible patients.

#### Criteria selection

The selection criteria for patient inclusion in our retrospective study were:

I. Adults (≥18 years) with a histopathological diagnosis of GB were included. While aiming to align with the WHO CNS5 (2021) classification, the retrospective nature of the cohort—collected prior to routine molecular profiling—limited the availability of molecular data. To ensure consistency, IDH1-mutant cases were excluded, and in the absence of molecular data, inclusion was based on MRI features characteristic of IDH1-wildtype GB (e.g., contrast enhancement, necrosis, peritumoral edema)^19^. Given the low prevalence of IDH1 mutations (∼5–10%) and the predominance of wildtype cases in pre-2021 cohorts, this approach was deemed appropriate to approximate the current WHO definition^20,21^.
II. A minimum survival period of 30 days post-surgery.
III. Total, supra-total or sub-total surgical resection, excluding biopsies or non-operable cases.
IV. Availability of essential MRI studies obtained by 1.5-T or 3-T scanners, depending on the specific purpose of each section. This includes morphological images: pre-and post-gadolinium T1-weighted (T1 and T1c), T2, FLAIR images, and, when available, perfusion information from dynamic susceptibility contrast MRI.

### Glioblastoma segmentation

Initially, the delineation of GB lesions was performed using the anatomical MRIs, categorizing them into three distinct tissues: I) CET, II) edema, and III) necrosis. This procedure includes preliminary preprocessing of the MRI scans, subsequently followed by segmentation employing convolutional neural networks (CNNs):

#### MRI preprocessing

MRI preprocessing involved several automated steps: all sequences were resampled to 1 mm^3^ isotropic voxels via linear interpolation, denoised using adaptive non-local means filtering ^22^, and rigidly registered within each patient to the contrast-enhanced T1-weighted image using ANTs ^23^. Subsequently, images were affine-transformed to MNI space ^24^ (ANTs), brain-extracted via a U-Net CNN pipeline, and corrected for magnetic field inhomogeneity with N4 ^25^ bias-field correction.

#### Lesion segmentation

A U-Net CNN model ^26^ processes 3D patches (32^3^ voxels) from T1c, T2, and FLAIR images across five hierarchical levels. Each level features four basic blocks (conv–batch norm–ReLU) followed by four residual blocks (adding a conv–batch norm–ReLU with skip connections). Filters increase from 16 to 256 with 3×3×3 kernels, incorporating down-and upsampling for multi-scale feature capture. Trained on BraTS 15, it achieved median Dice scores of 0.85 for contrast-enhancing tumor and 0.92 for total tumor, using L2 regularization and balanced batches to reduce overfitting and class imbalance ^26^.

### Non-contrast-enhancing tumor definition

The subsequent step involved segmenting the edema into two subregions: I) nCET and II) vasogenic edema. This segmentation was based on the intensity variations observed on T2 and FLAIR MRIs, following the principles outlined by Lasocki *et al^9^.* Specifically, the nCET region is defined as having relatively mild intensity on FLAIR and hyperintensity on T2, which may reflect its higher cellular density, as previous studies have shown an inverse correlation between cell density and FLAIR signal intensity^27^. In contrast, vasogenic edema is hyperintense on both sequences ^8,9^.

Preprocessed T2 and FLAIR MRIs were employed as inputs to execute the segmentation of the nCET. The morphological segmentation achieved in the preceding step was used, in which the FLAIR hyperintense region, including the nCET, is all classified as edema. The proposed methodology developed for the nCET segmentation task employs unsupervised Non-local Spatially Varying Finite Mixture Models^28^ (NLSVFMM). Specifically, this algorithm combines two techniques, Spatially Varying Finite Mixture Models (SVFMMs) and the Non-Local Means (NLM) framework, to accurately delineate the nCET region, while taking into account the anisotropy of the local features of the region.

Following NLSVFMM segmentation, we enforced three anatomical rules: (1) clusters must be spatially connected; (2) voxels >10 mm from the contrast-enhancing tumor (CET) were relabeled as edema; and (3) nCET regions touching the tumor core were dilated up to 10 mm ^9^ (in 1 mm steps), with any dilated regions losing core contact afterward reclassified as edema. This ensures physiologically plausible nCET delineations.

Following the applied constraints, to statistically confirm differences in intensity values between the identified nCET region and the remaining vasogenic edema, a Mann-Whitney U test was applied, considering results with p < 0.01 as significant. Additionally, to ensure that these differences were practically relevant, we introduced a secondary criterion based on the relative intensity difference. Specifically, we employed *Equation* (1), where *I*_*nCET*_represents the intensity values within the nCET region, and *I*_*Vasog. Edema*_represents the intensity values within the vasogenic edema region. This metric calculates the percentage difference between nCET and vasogenic edema intensities, normalized by the overall intensity range of the entire brain:

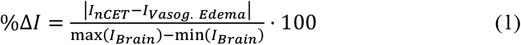

To be deemed practically significant, %Δ*I* must exceed 5%.

Following the segmentation of the nCET region, we investigated its relationship with the CET by analyzing two volumetric biomarkers: the ratio of CET to edema volume (CET/edema), and the ratio of nCET to edema volume (nCET/edema). These metrics reflect the spatial and structural interplay between distinct tumor compartments. Moreover, the size of CET and its resection conforms a hallmark of GB that has been extensively linked to patient prognosis^12^. To assess the strength and nature of these associations, we computed the Pearson correlation coefficient between each biomarker and the respective volumetric features, evaluating the presence of linear dependencies.

### Definition of the Diffuse Infiltration Index (DII)

To quantify the relevance of the nCET region for specific patients, we defined a novel biomarker derived from its volumetric properties, named the Diffuse Infiltration Index (DII). This reflects the proportion of the total peritumoral edema volume occupied by the nCET, highlighting its diffuse infiltrative nature beyond the CET boundaries.

To compute the DII, we obtained the volumetric measurements of the nCET and the total edema. The DII was then defined as the ratio of the nCET volume (*V*_*nCET*_) to the total edema volume (*V*_*edema*_), as shown in *Equation (2).*

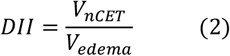

*Figure 1* illustrates the proposed pipeline for nCET segmentation and DII extraction, including the preprocessing of MRI scans, anatomical segmentation using the ONCOhabitats U-Net-based approach, edema subsegmentation with the NLSVFM model, and final computation of the DII.

**Fig. 1.**
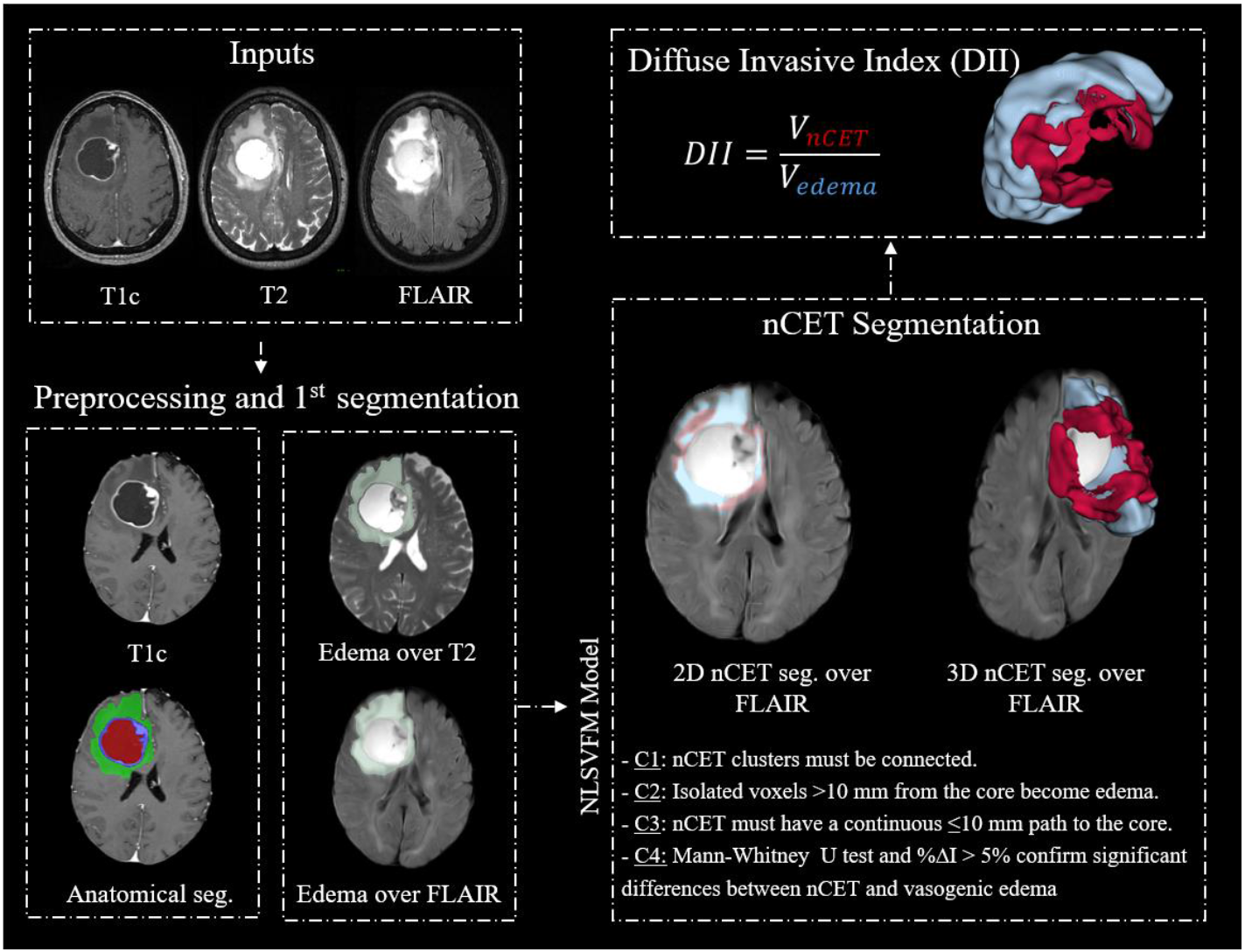
Graphical representation of the pipeline implemented for nCET segmentation and DII extraction. First, the input MR images are processed, and the anatomical segmentation is obtained using a U-NET-based approach ^26^ within the ONCOhabitats pipeline. This segmentation differentiates necrosis (red), CET (purple), and edema (green). Next, the NLSVFM model is applied to distinguish between vasogenic edema (blue) and nCET (red) within the segmented edema region, incorporating statistical and practical validations through predefined constraints. Finally, the DII was computed, quantitatively characterizing tumor infiltration and invasive heterogeneity.

### Assessment of DII prognostic capabilities

We assessed the prognostic capacity of the DII biomarker by evaluating its association with OS through univariate and multivariate cox regression analyses.

#### Univariate analysis

The association between the DII and OS was analyzed using univariate Cox regression, aiming to determine an optimal threshold that maximizes the concordance index (c-index) for stratifying patients into high- and low-survival groups. Subsequently, Kaplan-Meier curves were introduced to visually assess differences in survival probability distributions between the groups. The hazard ratio and p-value were also computed to quantify these differences.

In parallel, the same analytical approach was applied to the ratio between the volume of the CET and edema (CET/Edema), a widely used imaging-derived biomarker with well-established prognostic value^12^. Whereas CET/Edema captures the bulk of the enhancing tumor core—tissue that is typically removed during surgery—the DII quantifies the non-enhancing, infiltrative compartment that often remains after resection. Evaluating both biomarkers with identical methodology therefore allows a direct, like-for-like comparison between a conventional surgical target and our newly proposed infiltrative target, clarifying the additional prognostic information that DII may offer for pre-operative risk stratification and extent-of-resection planning.

Both the DII and the CET/Edema volume ratio were evaluated using data from the multi-center studies NCT03439332, GLIOCAT & BraTS, and OUS. These analyses were performed both independently for each dataset, and replicated using a combined, aggregated dataset.

#### Multivariate analysis

A multivariate Cox regression analysis was conducted to determine whether the prognostic influence of the DII on OS remained independent of routinely employed clinical variables. This analysis incorporated available clinical information, as well as volumetric and perfusion variables from MRI data part of the multicentric study NCT03439332 and GLIOCAT. The BraTS and OUS data were not included in this analysis because of missing clinical and perfusion data.

Clinical variables (gender, age, MGMT status) and imaging features (edema volume, 90th-percentile relative cerebral blood volume (rCBV) in nCET, DII) were encoded numerically—MGMT methylated = 0, unmethylated = 1, missing imputed by prevalence—and continuous variables scaled to [0,1]. Multivariate Cox models were built incrementally (clinical → +volumetric → +functional) with and without DII to assess its added prognostic value. Additionally, univariate Cox analyses of nCET and edema volumes was conducted on each of these factors individually.

### Software

The MRI preprocessing and anatomical segmentation were performed using the libraries presented by Juan-Albarracín et al. ^26,29^. The preprocessing pipeline and the nCET segmentation approach are fully integrated and accessible through the nCET segmentation service available at https://www.oncohabitats.upv.es/non-contrast-enhancing-tumor-segmentation/. Finally, the statistical analyses were implemented in Python (v3.8).

## Results

### Study cohort

*Figure 2* presents the CONSORT diagram illustrating the patient selection process based on the predefined inclusion criteria, detailing the stepwise approach applied to construct the final study cohort and the challenges encountered when applying the defined pipeline.

**Fig. 2.**
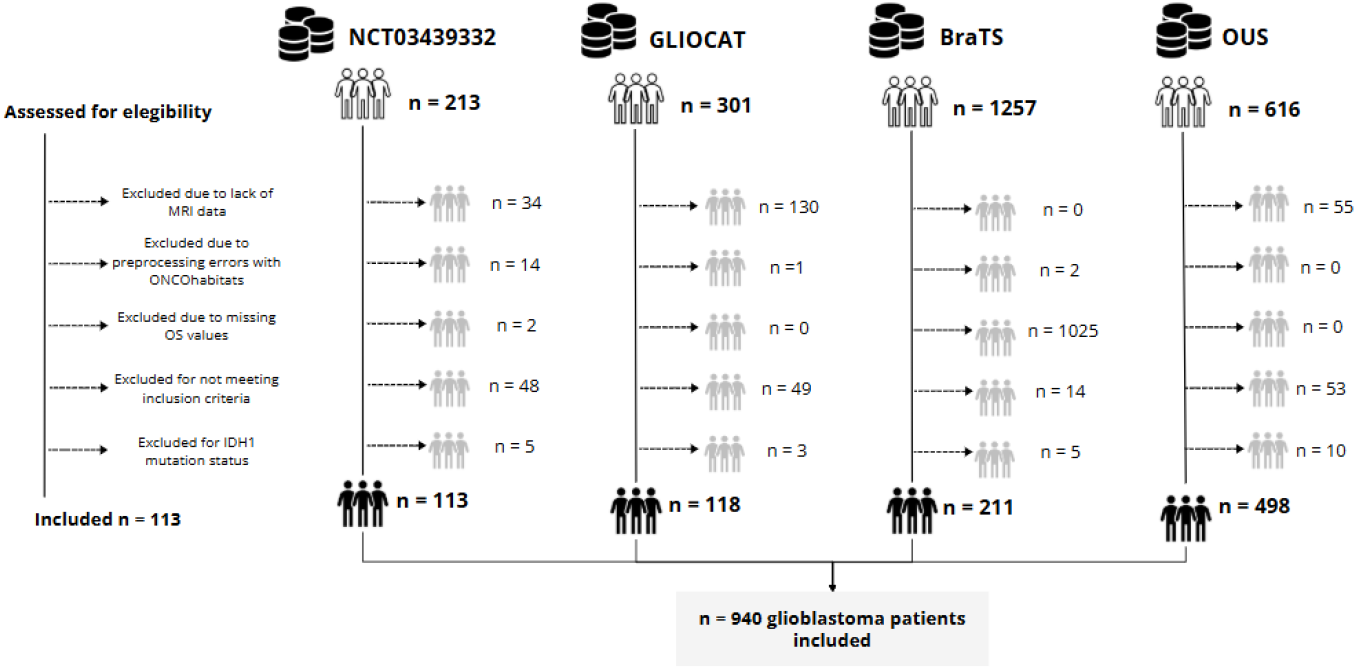
CONSORT diagrams for the NCT03439332, GLIOCAT, BraTS, and OUS datasets. Excluded patients with IDH1 mutations include those identified through molecular analysis, as well as those excluded based on radiological features not consistent with the IDH1-wildtype GB phenotype.

The final study cohort consisted of 940 patients diagnosed with glioblastoma, sourced from four datasets: 113 from NCT03439332, 118 from GLIOCAT, 211 from BraTS, and 498 from OUS. The median age (range) in years was 60 [35–81] for NCT03439332, 60 [32–80] for GLIOCAT, 62 [28–87] for BraTS, and 62 [28–87] for OUS. Median overall survival (range) in days was 406 [43–1229], 484 [70–2148], 386 [50–1767], and 411 [42–4919], respectively. Regarding survival status, 85 patients from NCT03439332 and 98 from GLIOCAT were deceased at the time of data collection, while survival data were not available for BraTS. In the OUS cohort, 482 patients had died and 26 were censored.

### Non-contrast-enhancing tumor definition

The segmentation of edema into two distinct regions yielded voxel clusters corresponding to the nCET and vasogenic edema. As depicted in *Figure 3A-B,* the LMSVFM algorithm was primed at distinguishing areas of mild image intensity in FLAIR and hyperintensity in T2, setting them apart from regions where both T2 and FLAIR sequences exhibited hyperintense values.

**Fig. 3.**
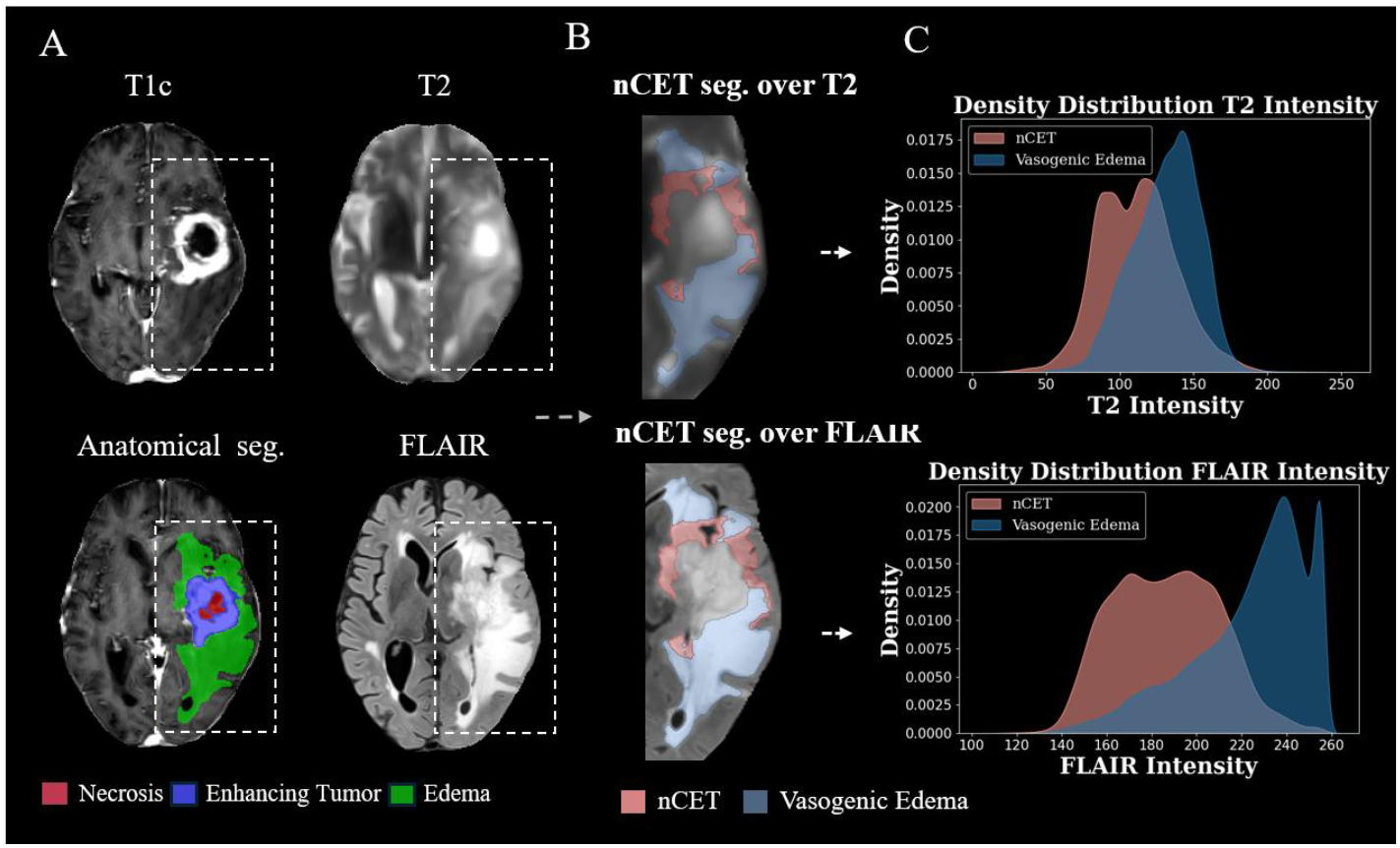
A) Results of nCET segmentation obtained from T2 and FLAIR MRIs, displayed alongside the anatomical tumor segmentation. B) Visualization of the segmented nCET and vasogenic edema regions overlaid on T2 and FLAIR sequences. C) Distribution of signal intensities in T2 and FLAIR for each region — example shown for patient OP03 from the NCT03439332 cohort.

Furthermore, the density distribution curves of intensity values in T2 and FLAIR in *Figure 3C* reveal a distinct pattern between the two regions. In the FLAIR sequence, nCET and vasogenic edema distributions exhibit notable heterogeneity, facilitating their differentiation. In contrast, the intensity distributions in T2 appear more homogeneous across both regions, leading to a high degree of overlap. This highlights the critical role of the FLAIR sequence in distinguishing nCET from vasogenic edema.

Mann-Whitney U tests for assessing the unsupervised segmentation revealed significant differences between the voxel distributions of the nCET and vasogenic edema across the study’s patient cohort in the three datasets: NCT03439332 (N = 113), Gliocat (N = 118), BraTS (N = 211), and BrainPower (N=498) with a total N = 940 (p-value< 0.01). The practical implications of this segmentation were evaluated by the %Δ*I*, as defined in *Equation* (1), revealing that for 917 out of 940 patients (97.5%) — 102 from the NCT03439332, 109 from Gliocat, 208 from BraTS and 498 from BrainPower — the segmentation achieved an intensity variation between the nCET and the vasogenic edema greater than 5%.

In the correlation analysis between the DII and the volumetric ratio of the CET to edema (CET/Edema), we observed that these two imaging-derived features are moderately correlated, with a Pearson correlation coefficient of r = 0.57. This results underscore that the CET, located within the tumor core and typically resected during surgery, is quantitatively associated with a peripheral region embedded in the edema — the nCET — automatically delineated through unsupervised segmentation.

### Assessment of DII prognostic capabilities

#### Univariate analysis

*Figure 4* provides an overview of the results from univariate analysis. Specifically, it presents the optimal thresholds for distinguishing between patients with high and low survival rates based on Cox regression, thereby establishing a link between nCET segmentation and OS. For the DII, the optimal threshold derived for each dataset (th = 0.71, 0.69, 0.69, 0.69) significantly differentiated the high-survival and low-survival groups (p < 0.05) with differences in median overall survival (OS) ranging from 62 to 122 days. At this threshold, the likelihood of mortality in the low-survival group was approximately twice as high as in the high-survival group in the NCT03439332 dataset (HR: 2.109 [95% CI: 1.33–3.25]; p = 0.0012). Notably, validating the threshold across independent datasets reinforces its robustness. In the GLIOCAT & BraTS and OUS datasets, while the differences in HR were less pronounced, they remained significant (HR: 1.31 [95% CI: 1.04–1.65]; p = 0.0217 and HR: 1.28 [95% CI: 1.07–1.53]; p = 0.0070, respectively). Finally, the analysis combining all datasets confirms the reliability of this partitioning, yielding an HR of 1.28 [95% CI: 1.12–1.47]; p = 0.0003 and establishing a definitive threshold of 0.69 for significantly distinguishing high-survival and low-survival groups. The corresponding difference in median overall survival—defined as the time point at which the estimated survival probability reaches 50%—between the two groups was 62 days.

**Fig. 4.**
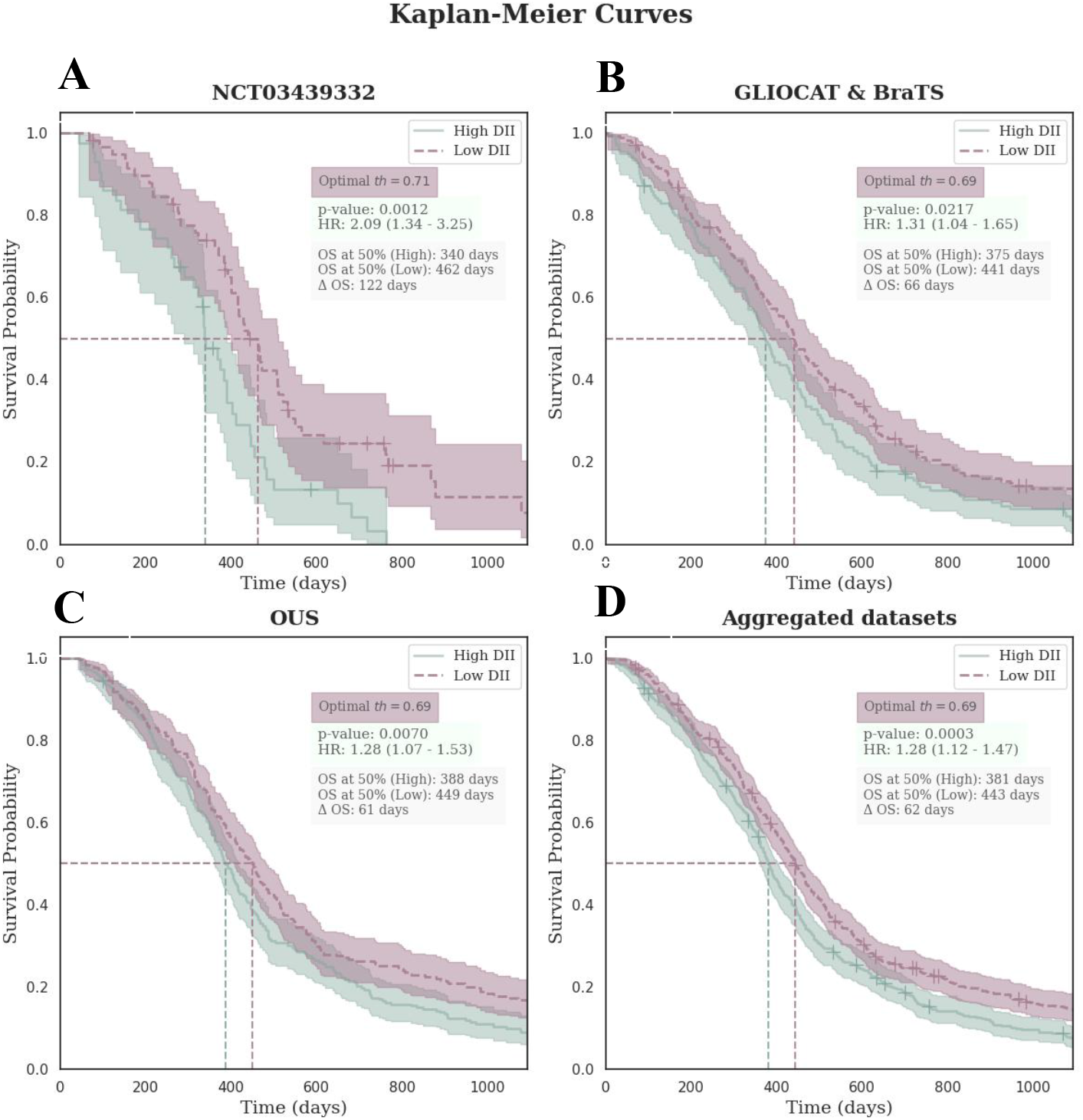
Kaplan-Meier curve for the optimal threshold obtained through the optimization of the C-index associated with Cox regression in NCT03439332, GLIOCAT & BraTS, OUS and all datasets combined for the DII, which biomarker separated high and low survivors. Additionally, the median survival time (when 50% of patients have died) was calculated for each group and the difference between them (Δ OS). To improve the visual interpretation of survival differences, all curves were truncated at 3 years (1095 days).

Moreover, the volumetric ratio of the enhancing tumor to edema (CET/edema) yielded significant associations with overall survival across all four experiments. The strongest result was observed in the NCT03439332 dataset (p = 0.0005, HR = 2.41 [1.47–3.98], ΔOS = 94), followed by the aggregated dataset (p = 0.0008, HR = 1.38 [1.14–1.68], ΔOS = 54). While the CET/edema ratio is correlated with overall survival—as expected given its established clinical relevance and the fact that the CET is routinely targeted during surgical resection—this variable was excluded from the final multivariate analysis, which specifically focused on the infiltrative nCET captured by the DII. Notably, the DII demonstrated a comparable prognostic performance to CET/edema, highlighting that the nCET region—although typically not completely resected—may carry equivalent prognostic value.

The Kaplan-Meier curves corresponding to the optimal threshold for each experiment in *Figure 4* show that more overall survival is expected for DII lower than the defined threshold. This difference is significant (p-value < 0.05). Moreover, analyzing the median survival time (mortality at 50%)—reveals a difference of 122 days for NCT03439332, 66 days for GLIOCAT & BraTS, 61 days for OUS, and 62 days across all datasets combined. This demonstrates that at the DII effectively differentiates two patient groups with an estimated survival gap of over two months.

#### Multivariate analysis

*Table 1* shows the effect of the different variables on OS based on Cox regression. The NCT03439332 and GLIOCAT datasets were used to determine if other variables already explain the DII prognostic relevance. Cox regression was first implemented including clinical variables, perfusion information and edema volume and it was noted that the significant variable is MGMT methylation status (HR: 1.92 [95% CI: 1.39 - 2.64]; p = 0.00006). In contrast, sex, age, perfusion and edema volume did not contribute to the model performance.

**Table 1.** Cox regression of clinical, perfusion and volumetric predictors (± DII) in NCT03439332 and GLIOCAT cohorts.

| Features | Excluding DII |  | Including DII |  |  |  |
| --- | --- | --- | --- | --- | --- | --- |
|  | CV + perfusion + volumetry |  | CV + perfusion |  | All variables |  |
|  | HR (95% CI) | p-value | HR (95% CI) | p-value | HR (95% CI) | p-value |
| Age | 1.22 [0.62–2.40] | 0.57 | 1.33 [0.67–2.62] | 0.41 | 1.35 [0.68–2.68] | 0.38 |
| Gender | 1.04 [0.77–1.40] | 0.82 | 1.03 [0.77–1.39] | 0.82 | 1.04 [0.77–1.40] | 0.81 |
| MGMT | 1.92 [1.39–2.64] | 0.00006<sup>b</sup> | 1.93 [1.41–2.64] | 0.00005<sup>b</sup> | 1.92 [1.40–2.63] | 0.00006<sup>b</sup> |
| rCBV<sub>max-nCET</sub> | 2.21 [0.73–6.65] | 0.16 | 2.31 [0.81–6.62] | 0.11 | 2.49 [0.85–7.35] | 0.09 |
| Edema volume | 0.73 [0.39–1.37] | 0.33 | - | - | 1.24 [0.57–2.69] | 0.58 |
| DII | - | - | 2.58 [1.22–5.47] | 0.013<sup>a</sup> | 3.00 [1.20–7.50] | 0.018<sup>a</sup> |
| C-index | 0.57 |  | 0.58 |  | 0.59 |  |

Abbreviations: CI = confidence interval; CV = clinical variables (age, gender, MGMT promoter methylation status); DII = diffuse infiltration index; HR = hazard ratio; nCET = non-contrast-enhancing tumor; rCBV = relative cerebral blood volume. <sup>a</sup> $p < 0.05$ ; <sup>b</sup> $p < 0.01$ .

When the DII was added to the same multivariate framework, its influence produces a significant contribution when analyzed with clinical variables and perfusion, and then including volumetric variables. The DII associated hazard ratio increases when volumetric information is added, implying that the edema volume enhance—but do not replace—the prognostic content of the DII; it still yields a significant p-value together with a larger effect size (HR: 3.00 [95% CI: 1.20 – 7.50]; p = 0.018). The methylation status of MGMT remains virtually unchanged across all experiments, suggesting that it does not represent a shared source of variation with the DII, edema volume, or perfusion metrics. Additionally, to verify that the information provided by the DII is not just related to any of its components alone, Cox regression was also performed on the volume of nCET and edema separately, showing that individually they are not significant (p>0.05).

*Figure 5* presents the results of the final multivariable Cox regression model, which includes sex, age, MGMT status, edema volume, perfusion, and the DII biomarker. The first panel displays the log HR, where significance is determined by whether the confidence interval crosses zero. A positive log HR indicates an increased risk (worse prognosis), while a negative log HR suggests a protective effect (better prognosis). The results show that MGMT methylation is associated with a better prognosis, as indicated by positive log HR. Moreover, DII analyzed as a continuous variable, shows a positive association with hazard, indicating that higher DII values correspond to a worse prognosis.

**Fig. 5.**
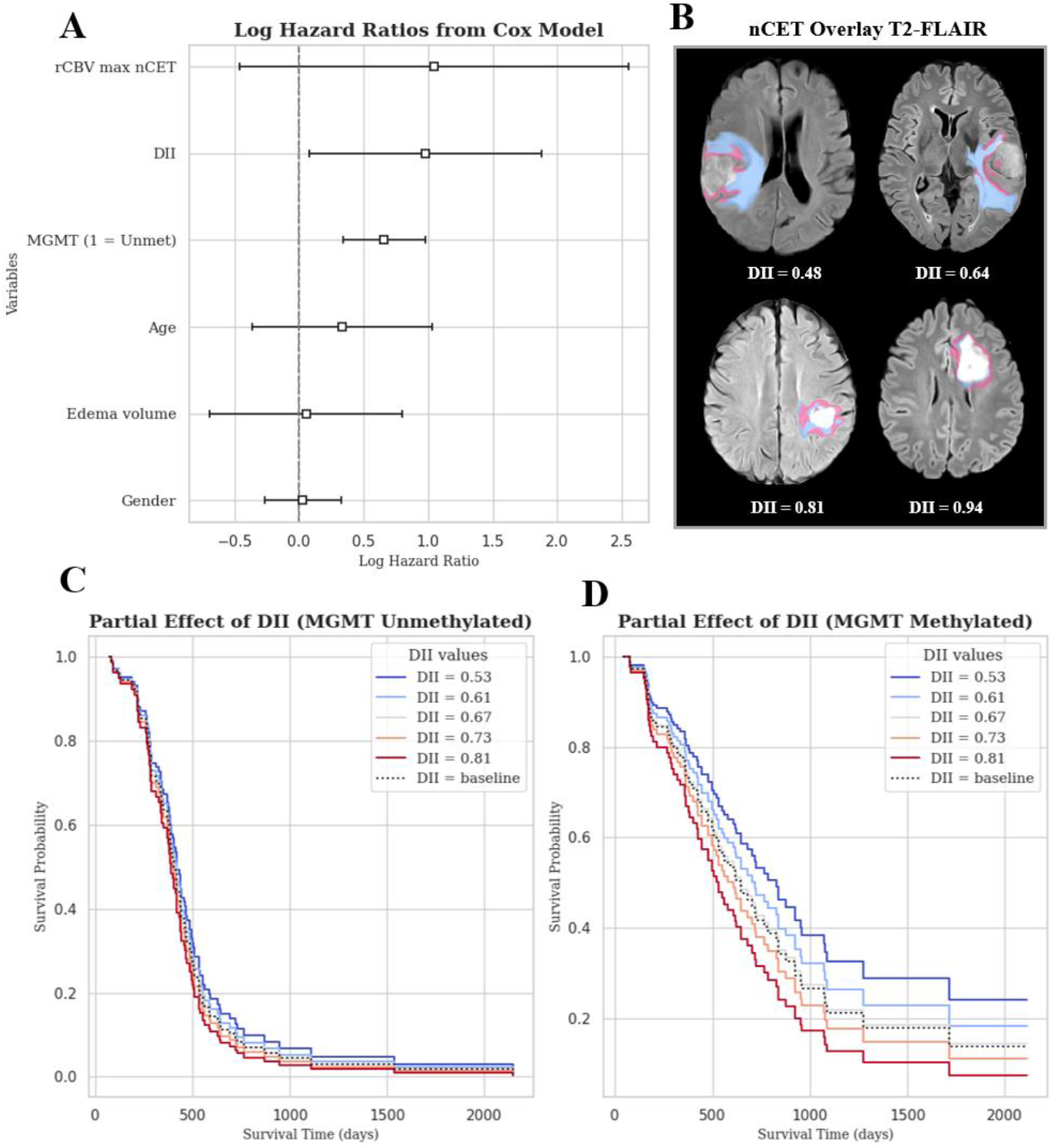
Cox proportional hazards model results and survival analysis based on DII for the last Cox Regression assessed (clinical variables, volumetric data, perfusion and DII). (A) Log HR from the Cox regression model, with the dashed line at 0 indicating the neutral effect threshold. Values to the left of the dashed line indicate a protective effect (lower risk), whereas values to the right indicate an increased risk. (B) T2-FLAIR images from a representative subset of patients, illustrating the spectrum of DII values. Segmentations of nCET (in magenta) and vasogenic edema (in blue) are overlaid on the anatomical scans. (C) Partial and conditional effect curves for MGMT-unmethylated patients and (D) for MGMT-methylated patients, illustrating the relationship between the continuous DII variable and OS. Survival probabilities are estimated at representative DII values while holding other covariates constant. The dotted line corresponds to the baseline DII, defined as the mean value of the DII distribution across the cohort.

*Figure 5C-D* presents the partial effect and conditional curves on OS as a function of the continuously analyzed DII variable, distinguishing between MGMT unmethylated (*C*) and methylated (*D*) patients. Notably, the model weights used in these curves correspond to those obtained in *Figure 5A*, reflecting the full-variable configuration to ensure that the partial effects of these variables are assessed within a validated framework, as indicated by the significance reported in the last row of *Table 1*. In this analysis, we observe that for MGMT-unmethylated patients, the partial effect of DII is nearly negligible relative to the baseline model. Conversely, MGMT-methylated patients exhibit a higher probability of longer OS with lower DII values (represented by cooler colors in *Figure 5D*), highlighting the association between DII and their influence on OS throughout the survival curve. *Figure 5B* displays a set of representative T2-FLAIR images from patients with varying DII values, illustrating differences in the size of the nCET component across the DII spectrum.

## Discussion

Our study presents a reproducible methodology for segmenting the nCET region in GB patients, distinguishing it from vasogenic edema based on commonly accessible MRI morphological sequences. The method is implemented as a standalone module on the Oncohabitats server and is freely accessible for research use. We validated the clinical significance of the nCET region delineated by assessing the prognostic capabilities of a derived biomarker, DII, in an GB multicentric datasets.

The nCET region harbors infiltrative tumor cells despite being outside the contrast-enhancing area that defines the tumor core ^8,9^. Accurate segmentation of this region based on T2 and FLAIR MRIs holds potential for understanding the complex architecture of GB. Our results demonstrate that 97,5% of patients exhibited a practical intensity difference between nCET and vasogenic edema. Morphological consistency was observed, as nCET clusters often maintained a connection to the tumor core (CET and necrotic region), reflecting potential migratory pathways for diffuse tumor infiltration.

The proposed DII biomarker was significantly associated with overall survival, offering prognostic information beyond conventional volumetric, perfusion, and genetic parameters. In particular, DII captures the infiltrative component of nCET relative to edema — a region that may remain after surgical resection. Remarkably, the prognostic performance of the DII (i.e., the nCET/edema ratio) was found to be nearly equivalent to that of the enhancing tumor CET/edema ratio. This is clinically relevant, as the CET region is routinely targeted and resected, whereas nCET is generally partially left in situ. That a non-resected, infiltrative compartment demonstrates prognostic power comparable to a surgically removed region underscores the biological significance of nCET and highlights the DII’s potential to guide both outcome prediction and therapeutic decision-making.

Our study underscores the DII’s unique prognostic value and its potential to complement existing clinical tools by capturing a biologically distinct and clinically meaningful dimension of tumor infiltration. The consistency of the optimal threshold (*th* = 0.69) across independent datasets further supports its robustness. Stratification of patients based on this threshold consistently distinguished between groups with different survival outcomes of more than two months, in line with our hypothesis that a higher DII reflects a more diffusely infiltrative and aggressive tumor phenotype.

MGMT methylation status was identified as a key predictor of overall survival, in line with established evidence ^30^. Importantly, among patients with MGMT-methylated tumors — a group generally associated with more favorable outcomes — the extent of nCET diffusely invading the edema, as captured by the DII, emerged as a particularly relevant prognostic factor. This observation suggests that a high DII may denote a more aggressive and infiltrative tumor phenotype, even within a biologically favorable subgroup. The capacity of the DII to quantify this infiltrative component adds complementary prognostic information beyond conventional markers, such as CET volume, extent of resection, MGMT methylation status and IDH1 mutation status^2,12,20,30^, supporting its potential utility for patient stratification in clinical trials and for tailoring treatment strategies. Additionally, MGMT methylation retained significance in the multivariate model, further reinforcing their clinical relevance.

Despite these promising results, the primary limitation of our study is the lack of gold standard for nCET segmentation. While the radiographic definition of nCET was employed, the clinical significance of the segmentation was corroborated through its association with OS and multi Cox regression analyses. Future work will address this limitation through histopathological validation and further clinical studies.

From a clinical perspective, the findings could have relevant implications for surgical planning. Current guidelines advocate supramaximal resection to improve survival in GB patients ^7,31^. However, there is no consensus on the extent of resection, particularly concerning the edema surrounding the tumor core, with less than 5 cm^3^ of nCET needing to remain for the procedure to be considered supramaximal ^7^. The methodology proposed in this study offers a reproducible tool to identify infiltrative and invasive regions within the apparent edema, providing neurosurgeons with critical information to refine surgical strategies and potentially improve patient outcomes.

## Data Availability

The BraTS dataset analysed in this study is openly available from the Multimodal Brain Tumor Segmentation Challenge repository (https://www.med.upenn.edu/cbica/brats).
The remaining datasets (OUS, Gliocat, and the NCT03439332 multicentre cohort from the hospitals of Alzira, Clinic de Barcelona, Liege, Manises, Oslo, Parma and Vall d'Hebron) contain sensitive patient-level information and are owned by the respective institutions. These data are therefore not publicly available but can be provided by the individual centres on reasonable request and subject to the relevant institutional ethics approval.

## Funding

Spanish Ministry of Science, Innovation and Universities (FPU23/02431); European Research Council (758657-ImPRESS); Helse Sør-Øst Regional Health Authority (2021057, 2017073); Research Council of Norway (325971, 261984, 303249, 32543); Fundació La Marató de TV3 (665/C/2013); Instituto de Salud Carlos III (PI18/01062); This work was funded by Grant PID2021-127110OA-I00 (PROGRESS) funded by MCIN/AEI/ 10.13039/501100011033 and by ERDF; Generalitat Valenciana (INVEST/2022/298, CIAICO/2022/064); Agencia Valenciana de la Innovación (INNEST/2022/87); Universitat Politècnica de València (PAID-01-24).

### Acknowledgments

ChatGPT (OpenAI, San Francisco, CA, USA) and Grammarly (Grammarly Inc., San Francisco, CA, USA) were used solely to refine English grammar; these tools did not contribute to the scientific content.

## Authorship

Conceptualization: MGM, JMGG, EFG, KEE; Methodology: MGM, CLM, JGT, VMM, SFS, KEE, EFG; Data curation: MGM, CLM, JGT, VMM, SFS, EEMM, EOVM, CBQ, JP; Formal analysis: MGM, CLM, JGT, VMM; Visualization (figures and tables): MGM, CLM; Writing – original draft: MGM; Writing – review & editing: all authors; Supervision: JMGG, EFG, KEE; Approval of final version of the manuscript: all authors.

